# Media exposure and knowledge of the ovulation cycle among adolescent girls who experienced menarche in Ghana: Evidence from the 2022 Demographic and Health Survey

**DOI:** 10.1101/2025.11.17.25340459

**Authors:** Emmanuella Elorm Lartey, Desmond Klu, Matilda Aberese-Ako, Emmanuel Harris, Rukaya Dongu Kamaldeen

## Abstract

Knowledge of the ovulation cycle among adolescent girls who have experienced menarche plays a major role in overcoming misconceptions about the fertile window within their cycles, which limits their ability to make informed decisions about sexual activity and contraceptive use. Mass media have become a powerful tool for health promotion and information sharing, especially among young populations. Despite the widespread use of mass media for health promotion, reproductive health knowledge among adolescent girls remains understudied. Therefore, this study aimed to determine the association between media exposure and knowledge of the ovulation cycle among adolescent girls who experienced menarche in Ghana. Data were extracted from the 2022 Ghana Demographic and Health Survey (GDHS). The data resulted in a weighted sample of 2,609 adolescent girls (mean age = 16.9 years and standard deviation ± 1.42). Descriptive statistics were used to summarize the study findings. Multivariable logistic regression was used to analyze the predictors of knowledge of the ovulation cycle among adolescent girls. All associations were considered statistically significant at the 95% confidence level.

The level of good knowledge of the ovulation cycle was 27.1%. Adolescent girls who were married (aOR=2.17; CI:1.24-3.78), who were cohabiting (aOR=1.75; CI:1.01-3.04), who were more educated (aOR=3.90; CI: 1.18-12.86), who used modern contraceptive methods (aOR=1.60; CI: 1.06-2.41), who used traditional contraceptive methods (aOR=2.70; CI:1.54-4.73) and who intended to use contraceptives later (aOR=1.53; CI: 1.19-1.98) had significantly greater odds of having good knowledge of the ovulation cycle.

Knowledge of the ovulation cycle among adolescent girls who experienced menarche in Ghana is relatively limited, which may contribute to unintended pregnancies and unsafe abortions. Future media should be thoughtfully designed for young audiences and integrated into broader reproductive health initiatives, particularly for adolescent girls with low levels of education, who are formally married, and those who do not know any contraceptive methods.

## Introduction

Adolescence, as defined by the World Health Organization (WHO), spans from age 10 to 19 years and is a vital period marked by rapid physical, emotional, and social changes. During this time, young people begin to think more deeply, develop their sense of identity, and navigate changing relationships with family, friends, and society. They also start to gain independence and take on more responsibility as they approach adulthood. However, this stage has its own challenges. Many adolescents encounter obstacles such as limited access to high-quality education, few job opportunities, and various health risks, including early or unintended pregnancies, sexually transmitted infections, exposure to violence, and increased mental health issues such as anxiety and depression (1,2). Adolescents constitute a significant proportion of the world’s population, approximately 1.3 billion. The settings in which they grow up, both physically and socially, are rapidly changing due to urbanization, shifting cultural norms, and evolving trends in marriage age and premarital sexual behaviour. As many young people begin exploring sexual relationships in their later teenage years, they often face major challenges in obtaining the right health information, advice, and services they need to make safe and informed decisions about their reproductive health (3). An estimated 21 million girls aged 15 – 19 years and 2 million girls under 15 years become pregnant in low- and middle-income countries (LMICs) (4,5). South Asia and sub-Saharan Africa (SSA) recorded the highest prevalence. A systematic review and meta-analysis revealed that the prevalence of adolescent pregnancy in sub-Saharan Africa was 19.3%, with substantial regional variation ranging from 16% in Central Africa to 22% in East Africa (6).

Ghana faces significant challenges in adolescent reproductive health, with a pooled prevalence of adolescent pregnancy estimated at 15.4% and many unintended pregnancies (5). Inadequate knowledge of reproductive physiology, especially regarding the menstrual cycle and ovulation timing, contributes to these negative outcomes. Studies from SSA indicate that many adolescents lack basic knowledge of menstruation, with 37.3% reporting that they do not know about it (7). In West Africa, fewer than half of adolescent girls know about menstruation before menarche, and many have misconceptions about the fertile window within their cycles, limiting their ability to make informed decisions about sexual activity and contraceptive use (8). Ovulation occurs when the dominant follicle releases its egg from the ovary into the fallopian tube for potential fertilization, and it is regulated by a surge in luteinizing hormone. Knowing when a woman’s period starts, when she ovulates, and when her next period is due is important for reducing the risk of unwanted or unplanned pregnancy (9–11). Knowledge of the ovulation cycle among adolescent girls is influenced by factors such as educational level, age, wealth, and media exposure (9,12,13). Mass media, including television, radio, and newspapers, have become powerful tools for health promotion and information sharing, especially among young populations. In many LMICs, media serve as a main source of information, often reaching groups that face barriers to formal education or health services. The accessibility, broad reach, and relative anonymity of mass media make it especially suitable for delivering sensitive health information to adolescents, who may feel uncomfortable seeking such info face-to-face (14). The impact of mass media on improving health knowledge and behaviours has been well documented across various African contexts, with studies showing that exposure to radio, TV, and print media is significantly associated with reproductive behaviours (15). Media campaigns have successfully increased awareness and understanding of maternal health services, with exposure to radio and TV being linked to increased antenatal care visits (16). Campaigns across SSA utilize various platforms, including newspapers, TV, radio, magazines, social media, and billboards, as they can reach large audiences at relatively low costs. TV provides visual and audio content that can effectively communicate complex health information, whereas radio reaches remote and rural areas where other media might not be available. Although newspapers reach fewer adolescents, they offer detailed information that can be reviewed and remembered (17).

Despite the widespread use of media for health promotion, the specific relationship between exposure to different media types (television, radio, and newspapers) and reproductive health knowledge among adolescents remains understudied. While some studies have examined the role of media in promoting contraceptive knowledge and use, few studies have specifically investigated the association between media exposure and knowledge of the ovulation cycle among adolescent girls in Ghana. The findings of this study will inform scalable, accessible interventions that leverage existing media infrastructure to empower adolescent girls with the knowledge they need to protect their reproductive health and make informed decisions about their bodies and futures.

## Methods

### Study design and sampling

This study employed a cross-sectional design based on the 2022 Ghana Demographic and Health Survey (GDHS). The 2022 GDHS was conducted by the Ghana Statistical Service (GSS) and aims to provide data for monitoring the population and health situation in Ghana. It is the seventh Demographic and Health Survey carried out in Ghana. The 2022 GDHS employs a stratified two-stage cluster sampling method designed to produce representative results at the national level, as well as for urban and rural areas and each of the 16 administrative regions, for most Demographic and Health Survey (DHS) indicators. In the first stage, 618 target clusters were selected from the sampling frame via a probability proportional to size (PPS) strategy, separately for urban and rural areas within each region. Next, the number of targeted clusters was selected through an equal probability systematic random sampling method from the clusters chosen in the first phase for both urban and rural areas in each region. In the second stage, after selecting the clusters, household listing and map updating were conducted in all selected clusters to create a list of households for each cluster. This list served as the sampling frame for selecting households for interviews. The household listing was performed using tablet computers equipped with software provided by the DHS Program. Finally, a fixed number of 30 households per cluster were randomly chosen from the list for interviews (18).

### Sample size

In the 2022 GDHS, 15,014 women aged 15 – 49 years were interviewed, contributing to a 98% response rate (19). Our study focused on adolescent girls aged 15-19 years who experienced menarche at the time of the survey. Among the 15,014 women, 12,285 women were excluded from the study because they were above the age of 19 years. A total sample size of 2,682 adolescent girls aged 15-19 years was determined. The dataset was then weighted, resulting in a final weighted sample of 2,609 adolescent girls aged 15–19 years who had experienced their first menstrual flow at the time of the survey.

### Study variables

#### Outcome variable

The outcome variable of our study was knowledge of the ovulation cycle. In the 2022 GDHS, participants were asked, “*From one menstrual period to the next, are there certain days when a woman is more likely to become pregnant?*” those who responded yes were asked a follow-up question: “*Is this time just before her period begins, during her period, right after her period ended, or halfway between two periods?*”. Possible answers to the questions were ‘just before her period begins’, ‘during her period’, ‘right after her period has ended’, ‘halfway between two periods’, ‘other’, and ‘don’t know’. From the responses, a binary category variable measuring knowledge of the ovulation cycle was derived. All those who stated “halfway between periods” were classified as having good ovulation knowledge, whereas the remaining responses were classified as having poor ovulation knowledge.

#### Independent variables

##### Media exposure factor

The main predictor variable for this study was media exposure; the frequency of watching television (not at all, less than once a week, at least once a week), frequency of listening to the radio (not at all, less than once a week, at least once a week), and frequency of reading newspapers or magazines (not at all, less than once a week, at least once a week) were recategorized as index variables (“1” = access to all media, “2” = access to at least one media, and “3” = access to no media).

##### Sociodemographic factors

The covariates included sociodemographic factors such as educational level (no formal education, primary education, secondary education, or higher education), marital status (not married, married, cohabiting, or formally married), ethnicity (Akan, Ewe, Ga/Dangme, Guan, Mole-Dagbani, Grusi, Gurma, or Mande), religious affiliation (orthodox, Pentecostal/charismatic, other Christian, Islam, traditional/spiritualist, or no religion), and literacy (illiterate, semiliterate, literate).

##### Household factors

The household factors considered for this study were the sex of the household head (male, female), the age of the household head (15 – 34, 35 – 54, 55+), the size of the household (1 – 3 members, 4 – 6 members, 7+ members), and the household wealth index (poorest, poorer, middle, richer, richest).

##### Sexual and reproductive factors

Sexual and reproductive factors, such as contraceptive use and intentions (using modern methods, traditional methods, which are intended for later use, do not intend to be used later), contraceptive knowledge (knowledge of no method, knowledge of only traditional methods, knowledge of modern methods), and the number of sexual partners (no partner, one partner, two or more partners), were included.

### Data analysis

The data were analyzed using STATA version 17. The data were weighted to make them nationally representative and provide a better statistical estimate. Descriptive statistics such as frequencies and percentages were used. The Pearson chi-square test was used to determine the associations between adolescent ovulatory cycle knowledge and the predictor variables. The Variance Inflation Factor (VIF) was also used to test for multicollinearity between the predictor variables, which indicated no evidence of multicollinearity (mean VIF = 1.30, maximum VIF = 0.55, minimum VIF = 0.94). Binary logistic regression analysis was conducted to assess the statistical associations among media exposure, sociodemographic, household, sexual, and reproductive factors, and adolescent knowledge of the ovulatory cycle. Two models were estimated: Model I examined the unadjusted association between media exposure and knowledge of the ovulatory cycle, whereas Model II adjusted for the combined effect of media exposure and sociodemographic, household, and sexual and reproductive health covariates. All variables were considered statistically significant at the 95% confidence interval (*p < 0*.*05*).

## Results

### Respondents’ background characteristics

**Table 1** presents the sociodemographic characteristics of the respondents. The mean age of the respondents was 16.9 years (standard deviation ± 1.42). Among the 2,609 adolescents interviewed, the majority (80.9%) had attained secondary education, with only a few (3.5%) reporting having no formal education, and the majority (91.9%) of them were not married. Ethnically, almost half (47.1%) were Akan, followed by Mole-Dagbani (17.5%) and Ewe (12.9%). Christianity was the predominant religion, particularly Pentecostal/charismatic (43.1%), while 18.5% identified as Muslim. Most (67.7%) respondents were literate. Household-level characteristics revealed that more than half (53.9%) of the household heads were aged 35–54 years and were male (56.2%). Additionally, nearly half (49.2%) of the households comprised 4 to 6 members, with relatively even distributions across wealth quintiles, with the majority (24.2%) of the respondents belonging to the middle class. Media exposure was common, with most (76.2%) reporting access to at least one medium. Additionally, the majority (72.6%) reported having no sexual partner. More than half (51.2%) did not intend to use contraception, although one-third (32.1%) indicated future intention. Only 11.5% of them were using modern contraceptive methods at the time of the survey. Despite low uptake, knowledge of contraception was nearly universal, with the majority (97.6%) being aware of modern contraceptive methods.

**Table 1:** Sociodemographic characteristics of the respondents.

| **Variables** | **Weighted frequency**<br>**(N=2609)** | **Percent (%)** |
| --- | --- | --- |
| **Marital Status** |  |  |
| Not married | 2,397 | 91.89 |
| Married | 82 | 3.15 |
| Cohabiting | 111 | 4.26 |
| Formally married | 18 | 0.70 |
| **Education** |  |  |
| No formal education | 91 | 3.49 |
| Primary education | 379 | 14.54 |
| Secondary education | 2,110 | 80.87 |
| Higher education | 29 | 1.10 |
| **Ethnicity** |  |  |
| Akan | 1,230 | 47.14 |
| Ga/Dangme | 155 | 5.94 |
| Ewe | 337 | 12.93 |
| Guan | 86 | 3.30 |
| Mole-dagbani | 458 | 17.54 |
| Grusi | 97 | 3.71 |
| Gurma | 155 | 5.93 |
| Mande | 91 | 3.51 |
| **Religious Affiliation** |  |  |
| Orthodox | 560 | 21.47 |
| Pentecostal/charismatic | 1,125 | 43.14 |
| Other Christian | 382 | 14.66 |
| Islam | 482 | 18.46 |
| Traditional/spiritualist | 24 | 0.90 |
| No religion | 35 | 1.36 |
| **Literacy** |  |  |
| illiterate | 442 | 16.96 |
| Semi-literate | 400 | 15.35 |
| Literate | 1,766 | 67.69 |
| **Household factors** |  |  |
| **Age of Household Head** |  |  |
| 15-34 | 300 | 11.48 |
| 35-54 | 1,405 | 53.85 |
| 55+ | 904 | 34.67 |
| **Sex of Household Head** |  |  |
| Male | 1,467 | 56.23 |
| Female | 1,142 | 43.77 |
| **Size of Household Head** |  |  |
| 1-3 members | 543 | 20.81 |
| 4-6 members | 1,282 | 49.15 |
| 7+ members | 784 | 30.04 |
| **Household Wealth Index** |  |  |
| Poorest | 423 | 16.20 |
| Poorer | 553 | 21.19 |
| Middle | 631 | 24.20 |
| Richer | 512 | 19.64 |
| Richest | 490 | 18.77 |
| **Media exposure factor** |  |  |
| **Media accessibility** |  |  |
| Access to all media | 315 | 12.07 |
| At least one media | 1,987 | 76.16 |
| Access to no media | 307 | 11.76 |
| **Sexual and reproductive health factors** |  |  |
| **Number of sexual partners** |  |  |
| No partner | 1,893 | 72.57 |
| One partner | 653 | 25.05 |
| Two or more partners | 62 | 2.38 |
| **Contraceptive use** |  |  |
| Using modern method | 299 | 11.47 |
| Using traditional method | 135 | 5.17 |
| Intends to use later | 838 | 32.13 |
| Does not intend to use | 1,336 | 51.23 |
| **Contraceptive knowledge** |  |  |
| Knows no method | 55 | 2.10 |
| Knows only traditional method | 8 | 0.33 |
| Knows modern method | 2,545 | 97.58 |

Source: Authors' computation from the GDHS 2022

### Associations between sociodemographic, household, media exposure, sexual and reproductive health factors, and knowledge of the ovulation cycle among adolescent girls who experienced menarche in Ghana

The bivariate analysis revealed an association between knowledge of the ovulation cycle and both the main predictor variable and the covariates, which included sociodemographic, household, media exposure, and sexual and reproductive health factors. **Table 2** presents the distribution of ovulation knowledge according to the sociodemographic, household, media exposure, and sexual and reproductive health characteristics of the respondents. Significant associations were observed between ovulation knowledge and marital status (p<0.001), education (p<0.001), ethnicity (p<0.01), religion (p<0.001), literacy (p<0.001), the household wealth index (p<0.001), media accessibility (p<0.001), contraceptive use (p<0.001), and contraceptive knowledge (p<0.01).

**Table 2:**
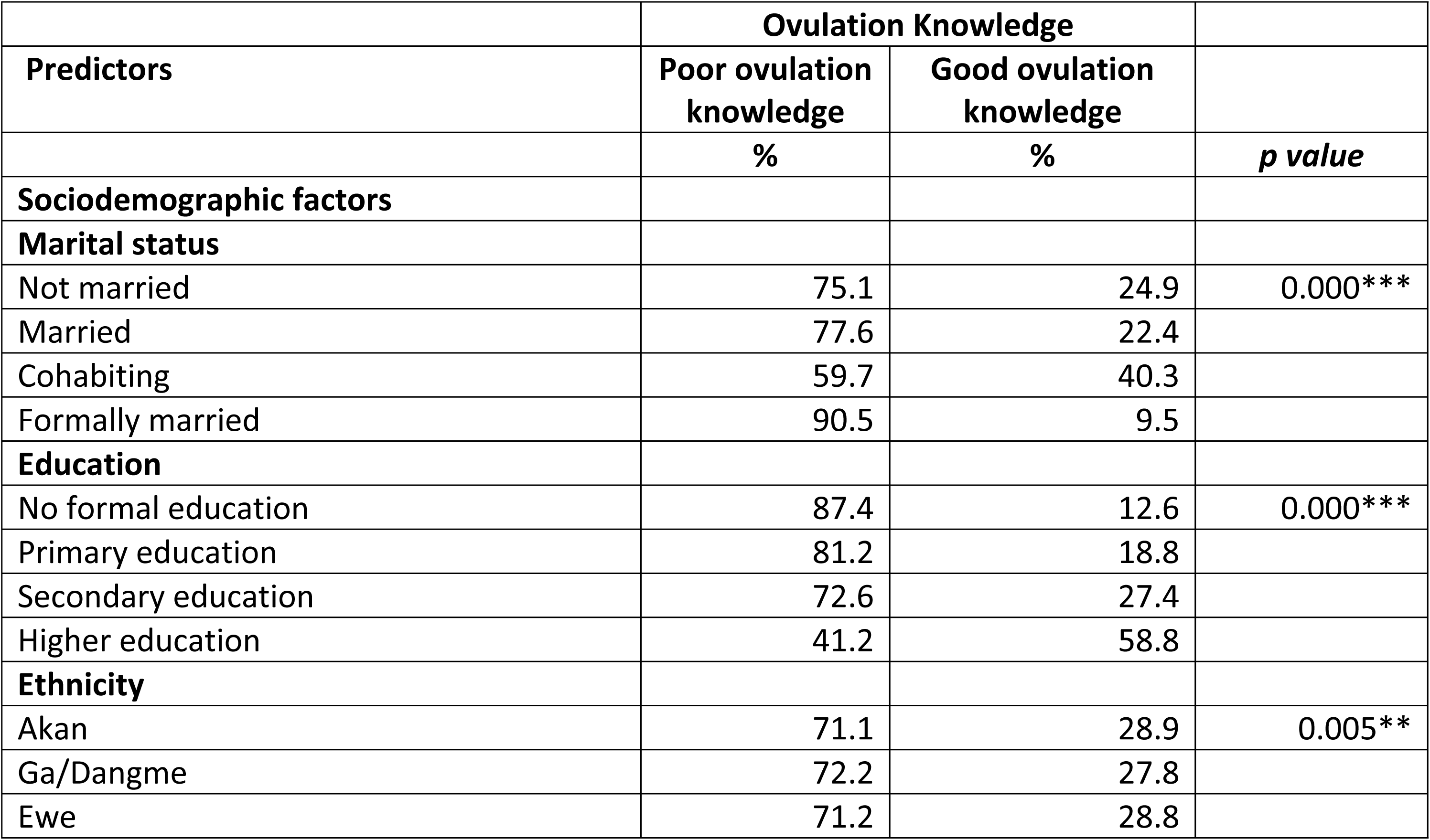
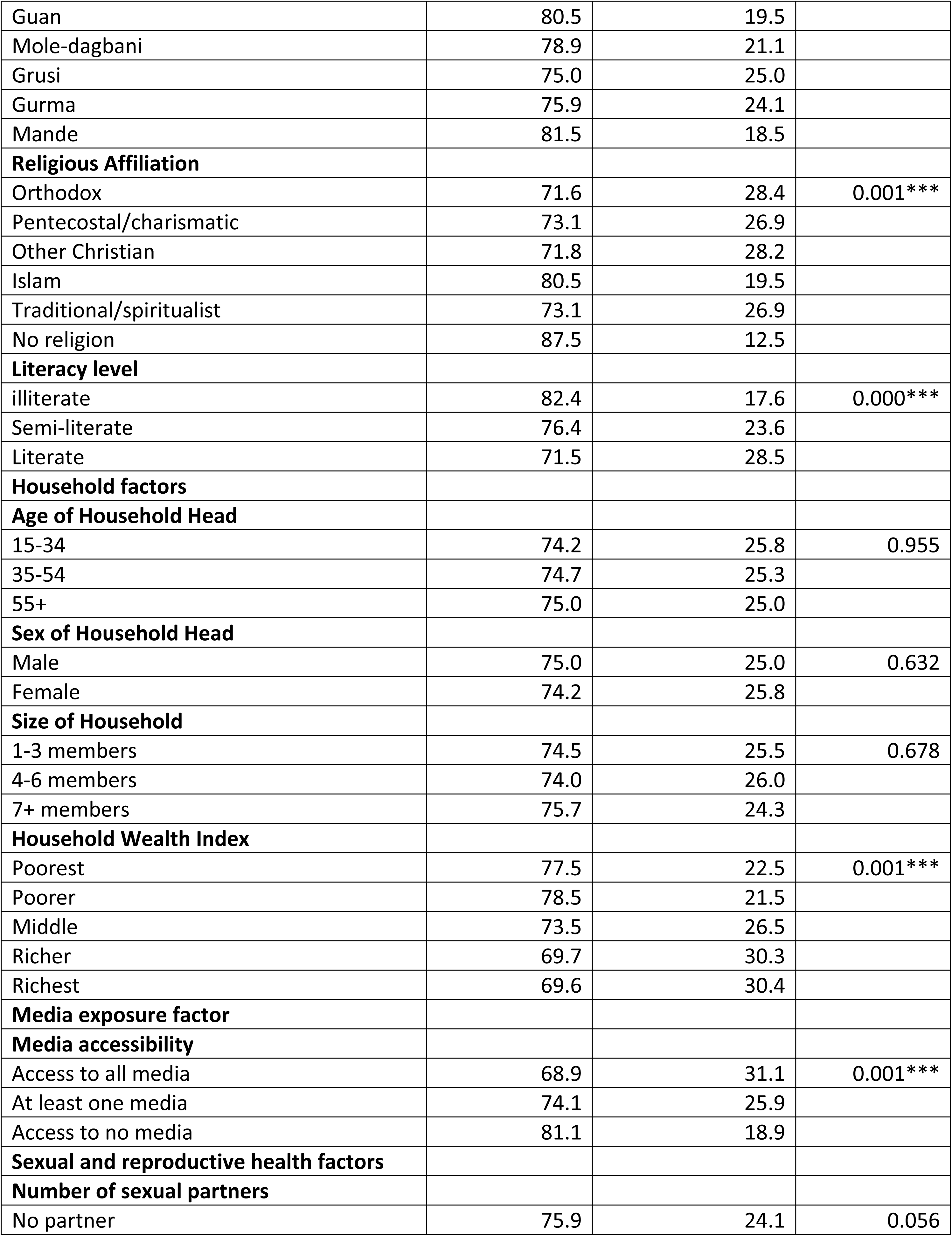
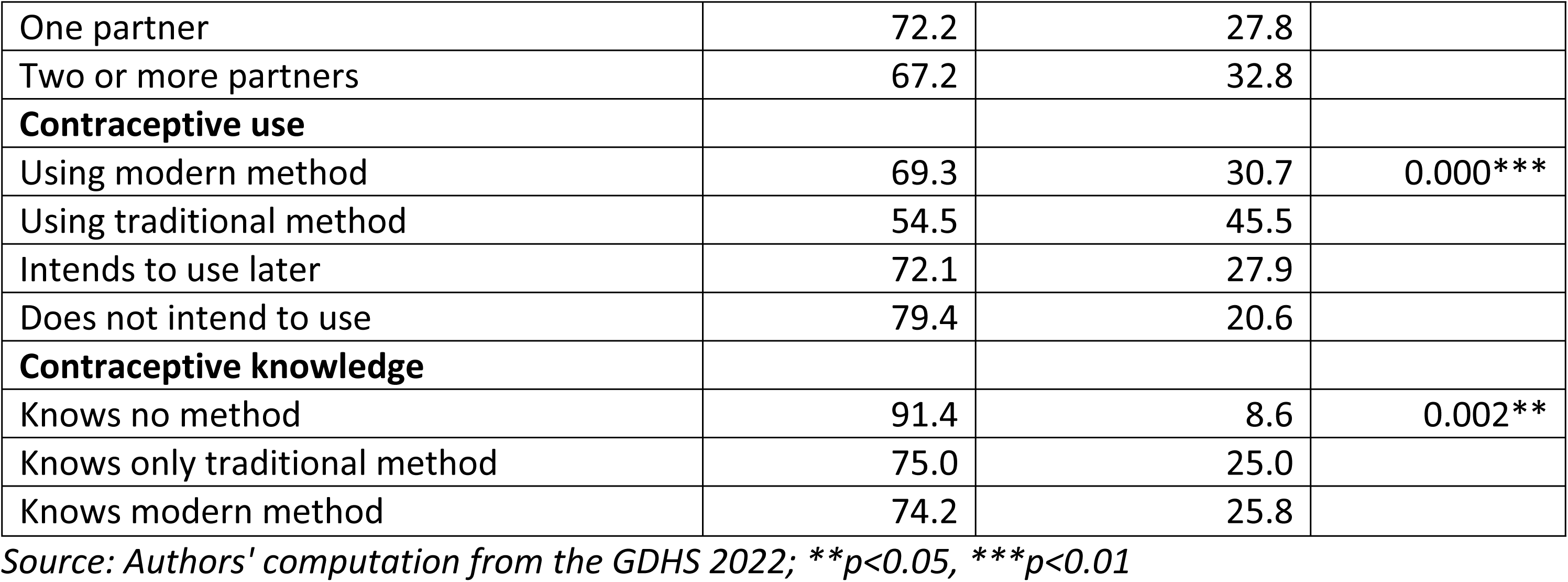
Association between predictor variables and knowledge of the ovulation cycle.

| Predictors | Ovulation Knowledge |  | *p value* |
| --- | --- | --- | --- |
|  | Poor ovulation knowledge | Good ovulation knowledge |  |
|  | % | % |  |
| **Sociodemographic factors** |  |  |  |
| **Marital status** |  |  |  |
| Not married | 75.1 | 24.9 | 0.000*** |
| Married | 77.6 | 22.4 |  |
| Cohabiting | 59.7 | 40.3 |  |
| Formally married | 90.5 | 9.5 |  |
| **Education** |  |  |  |
| No formal education | 87.4 | 12.6 | 0.000*** |
| Primary education | 81.2 | 18.8 |  |
| Secondary education | 72.6 | 27.4 |  |
| Higher education | 41.2 | 58.8 |  |
| **Ethnicity** |  |  |  |
| Akan | 71.1 | 28.9 | 0.005** |
| Ga/Dangme | 72.2 | 27.8 |  |
| Ewe | 71.2 | 28.8 |  |
| Guan | 80.5 | 19.5 |  |
| Mole-dagbani | 78.9 | 21.1 |  |
| Grusi | 75.0 | 25.0 |  |
| Gurma | 75.9 | 24.1 |  |
| Mande | 81.5 | 18.5 |  |
| **Religious Affiliation** |  |  |  |
| Orthodox | 71.6 | 28.4 | 0.001*** |
| Pentecostal/charismatic | 73.1 | 26.9 |  |
| Other Christian | 71.8 | 28.2 |  |
| Islam | 80.5 | 19.5 |  |
| Traditional/spiritualist | 73.1 | 26.9 |  |
| No religion | 87.5 | 12.5 |  |
| **Literacy level** |  |  |  |
| illiterate | 82.4 | 17.6 | 0.000*** |
| Semi-literate | 76.4 | 23.6 |  |
| Literate | 71.5 | 28.5 |  |
| **Household factors** |  |  |  |
| **Age of Household Head** |  |  |  |
| 15-34 | 74.2 | 25.8 | 0.955 |
| 35-54 | 74.7 | 25.3 |  |
| 55+ | 75.0 | 25.0 |  |
| **Sex of Household Head** |  |  |  |
| Male | 75.0 | 25.0 | 0.632 |
| Female | 74.2 | 25.8 |  |
| **Size of Household** |  |  |  |
| 1-3 members | 74.5 | 25.5 | 0.678 |
| 4-6 members | 74.0 | 26.0 |  |
| 7+ members | 75.7 | 24.3 |  |
| **Household Wealth Index** |  |  |  |
| Poorest | 77.5 | 22.5 | 0.001*** |
| Poorer | 78.5 | 21.5 |  |
| Middle | 73.5 | 26.5 |  |
| Richer | 69.7 | 30.3 |  |
| Richest | 69.6 | 30.4 |  |
| **Media exposure factor** |  |  |  |
| **Media accessibility** |  |  |  |
| Access to all media | 68.9 | 31.1 | 0.001*** |
| At least one media | 74.1 | 25.9 |  |
| Access to no media | 81.1 | 18.9 |  |
| **Sexual and reproductive health factors** |  |  |  |
| **Number of sexual partners** |  |  |  |
| No partner | 75.9 | 24.1 | 0.056 |
| One partner | 72.2 | 27.8 |  |
| Two or more partners | 67.2 | 32.8 |  |
| **Contraceptive use** |  |  |  |
| Using modern method | 69.3 | 30.7 | 0.000*** |
| Using traditional method | 54.5 | 45.5 |  |
| Intends to use later | 72.1 | 27.9 |  |
| Does not intend to use | 79.4 | 20.6 |  |
| **Contraceptive knowledge** |  |  |  |
| Knows no method | 91.4 | 8.6 | 0.002** |
| Knows only traditional method | 75.0 | 25.0 |  |
| Knows modern method | 74.2 | 25.8 |  |

Source: Authors' computation from the GDHS 2022; \*\* $p < 0.05$ , \*\*\* $p < 0.01$

The results further indicated that adolescents with higher education had the highest proportion (58.8%) of good knowledge of the ovulatory cycle compared with those with no formal education (12.6%). Similarly, literate respondents had greater ovulation knowledge (28.5%) than illiterate respondents did (17.6%). The highest proportion (40.3%) of adolescents who were cohabiting had good knowledge of the ovulatory cycle, and the lowest proportion (9.5%) were formally married. Comparatively, the respondents who had good ovulation knowledge belonged to the Akan tribe than those who belonged to the Mande tribe. Adolescents who were affiliated with the Orthodox church constituted the highest proportion (28.4%) of those with good ovulation knowledge, and the lowest proportion (12.5%) was observed among those with no religious affiliation. The highest proportion (30.4%) of the respondents who had good ovulation knowledge belonged to the richest category of wealth, while the lowest proportion (22.5%) belonged to the poorest category. Additionally, respondents with access to all media reported higher levels of good knowledge (31.1%) than those with no media access (18.9%). Compared with those not intending to use contraception (20.6%), adolescents using traditional contraceptive methods had greater ovulation knowledge (45.5%). In comparison, respondents who were familiar with modern contraceptive methods had greater ovulation knowledge (25.8%) than did those who were not (8.6%).

### Level of knowledge about the ovulatory cycle

With respect to the level of ovulatory cycle knowledge among adolescent girls who experienced menarche in Ghana, only 27.1% had good knowledge. *(See **Error! Reference source not found.**)*

**Fig 1:**
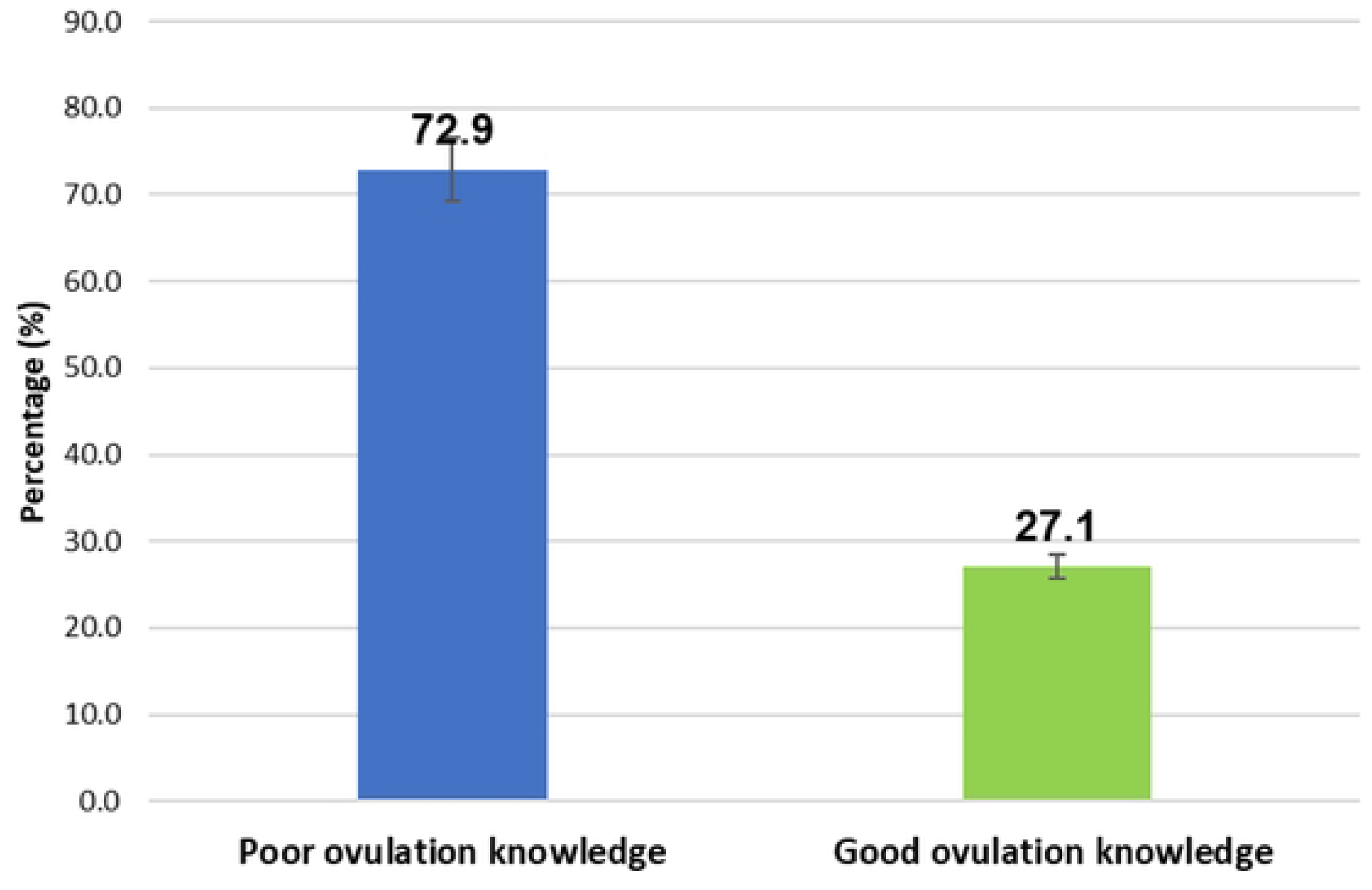
Level of knowledge about the ovulatory cycle among adolescent girls who experienced menarche.

### Predictors of knowledge of the ovulatory cycle among adolescent girls who experienced menarche

The second model (Model II) in ***Table 3*** revealed the combined effects of sociodemographic, household, media exposure, and sexual and reproductive factors on knowledge of the ovulatory cycle among adolescent girls who experienced menarche in Ghana. The findings indicate that marital status, education, literacy level, contraceptive use, and contraceptive knowledge were significantly associated with knowledge of the ovulatory cycle. Specifically, adolescent girls who were married were 2.17 times more likely to have good ovulatory cycle knowledge than their unmarried peers (aOR = 2.17; CI: 1.24–3.78). Additionally, those who were cohabiting had 1.75 times greater odds (aOR=1.75; CI: 1.01–3.04) of having good knowledge than those who were not married. However, adolescent girls who were formally married were 87% less likely (aOR=0.13; CI: 0.02–0.74) to have good ovulatory knowledge than their unmarried peers. Regarding education, respondents who had higher education had 3.9 times greater odds (aOR=3.90; CI: 1.18–12.86) of having good ovulatory cycle knowledge than those with secondary education. Compared with literate adolescent girls, illiterate adolescent girls had 46% lower odds (aOR=0.54; CI: 0.38–0.77) of having good ovulatory cycle knowledge. The results further indicate that adolescent girls using modern contraceptive methods were 1.6 times more likely (aOR =1.60; CI: 1.06–2.41) than nonusers. Additionally, those using traditional methods had 2.70 times higher odds (aOR = 2.70; CI: 1.54–4.73) than their nonpeer users. Adolescent girls who intended to use contraception later had 1.53 times higher odds (aOR= 1.53; CI: 1.19–1.98) than those who did not intend to use it. Finally, adolescent girls who reported knowing no contraceptive method were 76% less likely (aOR = 0.24; CI: 0.09–0.63) to have good ovulatory cycle knowledge than those who knew modern contraceptive methods.

**Table 3:**
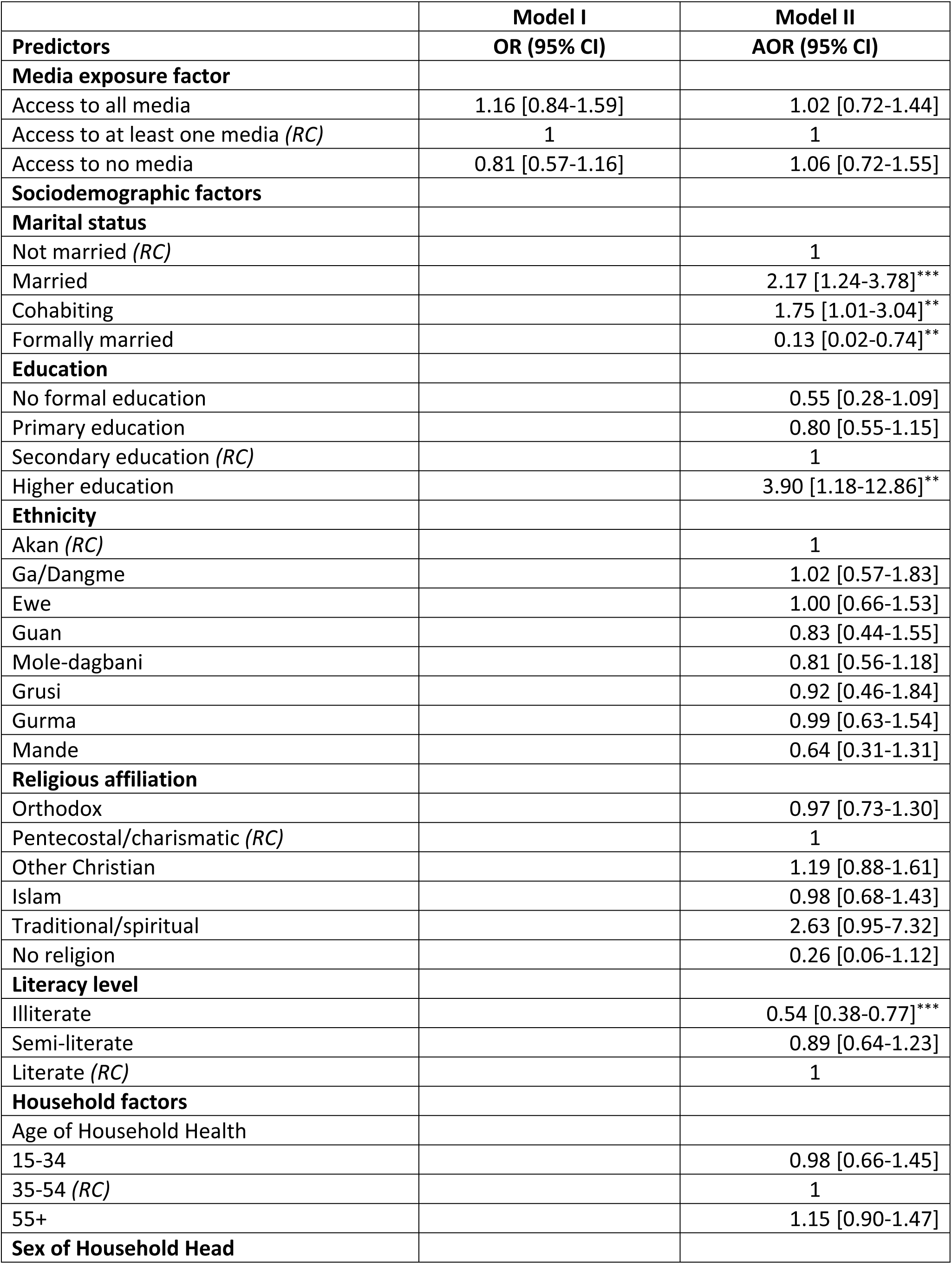
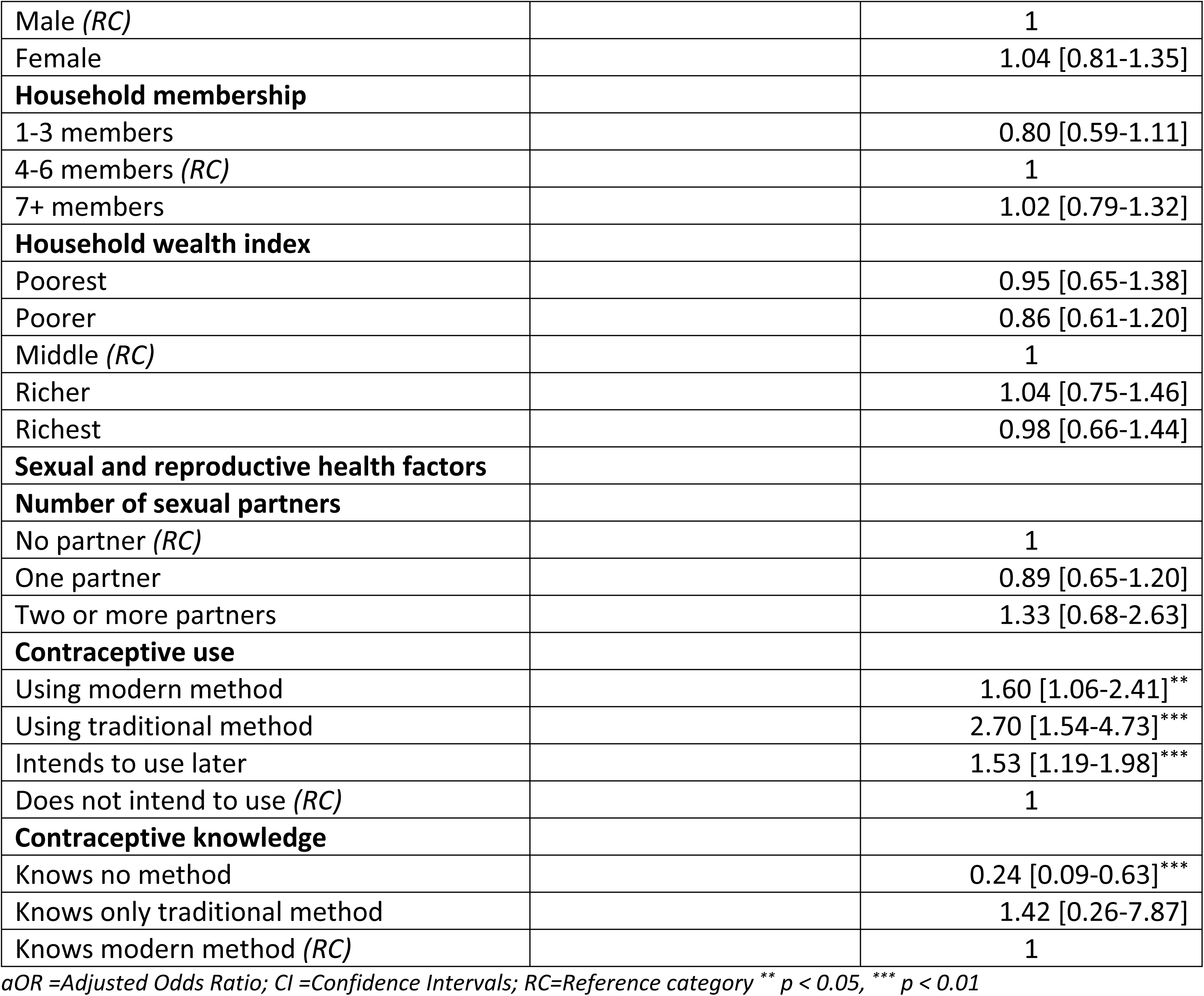
Binary logistics regression analysis of factors associated with knowledge of the ovulatory cycle.

| Predictors | OR (95% CI) | AOR (95% CI) |
| --- | --- | --- |
| **Media exposure factor** |  |  |
| Access to all media | 1.16 [0.84-1.59] | 1.02 [0.72-1.44] |
| Access to at least one media (RC) | 1 | 1 |
| Access to no media | 0.81 [0.57-1.16] | 1.06 [0.72-1.55] |
| **Sociodemographic factors** |  |  |
| **Marital status** |  |  |
| Not married (RC) |  | 1 |
| Married |  | 2.17 [1.24-3.78]*** |
| Cohabiting |  | 1.75 [1.01-3.04]** |
| Formally married |  | 0.13 [0.02-0.74]** |
| **Education** |  |  |
| No formal education |  | 0.55 [0.28-1.09] |
| Primary education |  | 0.80 [0.55-1.15] |
| Secondary education (RC) |  | 1 |
| Higher education |  | 3.90 [1.18-12.86]** |
| **Ethnicity** |  |  |
| Akan (RC) |  | 1 |
| Ga/Dangme |  | 1.02 [0.57-1.83] |
| Ewe |  | 1.00 [0.66-1.53] |
| Guan |  | 0.83 [0.44-1.55] |
| Mole-dagbani |  | 0.81 [0.56-1.18] |
| Grusi |  | 0.92 [0.46-1.84] |
| Gurma |  | 0.99 [0.63-1.54] |
| Mande |  | 0.64 [0.31-1.31] |
| **Religious affiliation** |  |  |
| Orthodox |  | 0.97 [0.73-1.30] |
| Pentecostal/charismatic (RC) |  | 1 |
| Other Christian |  | 1.19 [0.88-1.61] |
| Islam |  | 0.98 [0.68-1.43] |
| Traditional/spiritual |  | 2.63 [0.95-7.32] |
| No religion |  | 0.26 [0.06-1.12] |
| **Literacy level** |  |  |
| Illiterate |  | 0.54 [0.38-0.77]*** |
| Semi-literate |  | 0.89 [0.64-1.23] |
| Literate (RC) |  | 1 |
| **Household factors** |  |  |
| Age of Household Head |  |  |
| 15-34 |  | 0.98 [0.66-1.45] |
| 35-54 (RC) |  | 1 |
| 55+ |  | 1.15 [0.90-1.47] |
| **Sex of Household Head** |  |  |
| Male (RC) |  | 1 |
| Female |  | 1.04 [0.81-1.35] |
| **Household membership** |  |  |
| 1-3 members |  | 0.80 [0.59-1.11] |
| 4-6 members (RC) |  | 1 |
| 7+ members |  | 1.02 [0.79-1.32] |
| **Household wealth index** |  |  |
| Poorest |  | 0.95 [0.65-1.38] |
| Poorer |  | 0.86 [0.61-1.20] |
| Middle (RC) |  | 1 |
| Richer |  | 1.04 [0.75-1.46] |
| Richest |  | 0.98 [0.66-1.44] |
| **Sexual and reproductive health factors** |  |  |
| **Number of sexual partners** |  |  |
| No partner (RC) |  | 1 |
| One partner |  | 0.89 [0.65-1.20] |
| Two or more partners |  | 1.33 [0.68-2.63] |
| **Contraceptive use** |  |  |
| Using modern method |  | 1.60 [1.06-2.41]** |
| Using traditional method |  | 2.70 [1.54-4.73]*** |
| Intends to use later |  | 1.53 [1.19-1.98]*** |
| Does not intend to use (RC) |  | 1 |
| **Contraceptive knowledge** |  |  |
| Knows no method |  | 0.24 [0.09-0.63]*** |
| Knows only traditional method |  | 1.42 [0.26-7.87] |
| Knows modern method (RC) |  | 1 |

aOR =Adjusted Odds Ratio; CI =Confidence Intervals; RC=Reference category \*\* $p < 0.05$ , \*\*\* $p < 0.01$

## Discussion

This study used data from the 2022 GDHS to assess media exposure and knowledge of the ovulation cycle (KOC) among adolescent girls who experienced menarche in Ghana. The findings reveal that 27.1% of adolescent girls in Ghana have good KOC. This finding is much greater than what has been reported among adolescent females in Ghana (24.6%) (20). Similarly, lower rates of good KOC have been reported among women in Haiti (24.1%) and Ethiopia (23.6%) (21,22). However, the United States (32.8%), Togo (42.8%), and Cote d’Ivoire (30.4%) reported higher rates of good KOC (23,24). The difference in the prevalence of KOC might be due to the educational system, access to mass media, exposure, and the diverse study population. While a recent study was conducted among adolescent girls who experienced menarche in Ghana, Afrifa-Anane’s (20) study included all adolescent females, regardless of whether they had begun menstruating. Similarly, the studies of Jean Simon et al. in Haiti (21) and Ross et al. in Ethiopia (22) focused solely on reproductive-aged women. The disparities between adolescent girls and women may be due to peer networks and information sharing. Adolescent girls often discuss menstruation and reproductive health with their peers, especially after menarche, whereas adult women dispersed across households and workplaces may have fewer such networks.

Our multivariate analysis revealed that marital status, educational level, literacy level, contraceptive use, and contraceptive knowledge were significantly associated with good KOC among adolescent girls who experienced menarche. Our study revealed that adolescent girls who experienced menarche had substantially greater odds of having KOC than their unmarried counterparts. This finding is similar to that of a study conducted across 29 sub-Saharan African countries, which revealed that married young women who did not have correct KOC were more likely to have an unintentional pregnancy than never-married or previously married women (25). This finding may be as result of married adolescent girls having immediate and practical reasons to understand their fertility and to achieve or avoid pregnancy compared to their unmarried counterparts. The likelihood of KOC among adolescent girls who experienced menarche increased significantly with increasing educational status, which is consistent with findings from previous studies (26,27). Education may impart feelings of self-worth and self-confidence, which are necessary for changing health behaviour and seeking out health services. Structured curriculum delivery may ensure comprehensive coverage of topics, including menstrual cycles, ovulation, and fertility. Our study revealed that illiterate adolescent girls had lower odds of having good KOC than literate adolescent girls. This finding is similar to those of studies conducted in Nigeria (28) and Tanzania (29). A key reason why literacy affects KOC may be the inability to access written health information. Illiterate adolescents are unable to read reproductive health pamphlets, brochures, educational materials distributed in clinics or schools; health articles in newspapers or magazines, and even understand text messages containing health information. This severely limits their exposure to accurate reproductive health information that their literate peers can easily access. Adolescent girls who reported knowledge of contraceptives and the current use of contraceptive methods were associated with correct KOC, which is similar to the findings of a study conducted in Malawi (30). Access to modern or traditional methods, or even the intention to use contraception, appears to foster reproductive health literacy. These girls are likely to receive counselling or education when accessing modern methods, which may improve their understanding of fertility and ovulation. Traditional method users may reflect cultural or peer-driven knowledge sharing, where traditional practices are passed down with explanations of timing and fertility, and girls who intend to use contraception later could indicate proactive learning or exposure to reproductive health information even before actual use begins.

## Limitations

The study used data from the GDHS, and because the dataset is cross-sectional, it cannot establish causal relationships. While this study relies on nationally representative data, the findings may not be fully applicable outside the Ghanaian context. Cultural norms, media access, and reproductive health education differ across countries, so the patterns observed might look different elsewhere. Additionally, knowledge of the ovulation cycle was self-reported, which could introduce recall bias, as adolescent girls might underreport sensitive questions. Despite these limitations, this study may be one of the few in SSA that used nationally representative data to examine media exposure and knowledge of the ovulation cycle among adolescent girls who experienced menarche. Therefore, the findings contribute to the adolescent reproductive health literature in Ghana and SSA overall.

## Conclusion

This study revealed a low level of KOC among adolescent girls who experienced menarche in Ghana. Interestingly, mass media exposure, our main predictor of interest, was not significantly linked to knowledge of the ovulatory cycle among adolescent girls after we adjusted for sociodemographic, household, sexual, and reproductive health factors. Several plausible reasons exist for this finding. First, the measurement of media exposure in this study captured general media use rather than specific exposure to information about the ovulatory cycle. Adolescents may have regular access to media but limited exposure to programming that discusses fertility awareness. Second, once education and literacy, which are strong predictors of both media access and health knowledge, were controlled for in the multivariable model, media exposure did not provide additional explanatory power. Although the association with mass media was not statistically significant, this does not mean that mass media has no role. Instead, the findings suggest that media exposure alone is not enough. To enhance effectiveness, it should be paired with other approaches, such as comprehensive sex education in schools and communities, peer-led programs, and youth-friendly health services, especially those that focus on ovulatory cycle knowledge. Understanding the ovulatory cycle is key to preventing unintended pregnancies and unsafe abortions. Therefore, future media campaigns should be thoughtfully designed for young audiences with age-appropriate content and integrated into broader reproductive health initiatives.

## Data Availability

The data used for this study is publicly available from the Demographic and Health Survey dataset and be accessible via: https://dhsprogram.com/data/available- datasets.cfm

https://dhsprogram.com/data/available-datasets.cfm

## Acknowledgements

The authors thank the MEASURE Demographic and Health Survey program (DHS) and ICF International for granting access to the dataset for this study.

## Ethical approval

The Ghana Demographic and Health Survey is publicly available, and for this study, no ethical clearance was required. However, before conducting the analysis, permission and request were obtained from the MEASURE Demographic and Health Survey program.

## Supporting information

**S1 Table. Multicollinearity test using the Variance Inflation Factor (VIF)**

## References

1. Blum RW, Mmari K, Moreau C. It Begins at 10: How Gender Expectations Shape Early Adolescence Around the World. Journal of Adolescent Health. 2017 Oct 1;61(4):S3–4.

2. Singh JA, Siddiqi M, Parameshwar P, Chandra-Mouli V. World Health Organization Guidance on Ethical Considerations in Planning and Reviewing Research Studies on Sexual and Reproductive Health in Adolescents. Journal of Adolescent Health. 2019 Apr 1;64(4):427–9.

3. Lohan M, Brennan-Wilson A, Tomlinson M, of History S, Bradshaw MM, Child N, et al. School of Nursing and Midwifery ( Global research priority-setting exercise on the sexual and reproductive health and rights of young adolescents. Lancet Child Adolesc Health [Internet]. 2025;9:724–34. Available from: www.thelancet.com/child-adolescent

4. Tresha Lalla-Edward S, Chandiwana N, Maharaj NR. EDITED BY REVIEWED BY Adolescent pregnancy in sub-Saharan Africa-a cause for concern. 2022;

5. Mohammed S. Analysis of national and subnational prevalence of adolescent pregnancy and changes in the associated sexual behaviours and sociodemographic determinants across three decades in Ghana, 1988-2019. BMJ Open [Internet]. 2023;13:14. Available from: 10.1136/bmjopen-2022-068117

6. Kassa GM, Arowojolu AO, Odukogbe AA, Yalew AW. Prevalence and determinants of adolescent pregnancy in Africa: a systematic review and Meta-analysis. Reprod Health. 2018 Dec 29;15(1):195.

7. Finlay JE, Assefa N, Mwanyika-Sando M, Dessie Y, Harling G, Njau T, et al. Sexual and reproductive health knowledge among adolescents in eight sites across sub-Saharan Africa. Tropical Medicine & International Health. 2020 Jan 8;25(1):44–53.

8. Tomlinson MM, Wallis AB, Harris MJ, DuPré NC, Baumgartner RN, Okonofua F. Menstrual hygiene management among adolescent girls in West Africa: A systematic review. Afr J Reprod Health. 2024 Jan 31;28(1):123–56.

9. Diress G, Ahmed M, Adane S, Linger M, Alemnew B. Barriers and Facilitators for HIV Testing Practice Among Ethiopian Women Aged 15-24 years: Analysis of the 2016 Ethiopian Demographic and Health Survey. HIV/AIDS - Research and Palliative Care. 2021 Jan;Volume 12:963–70.

10. Owen M. Physiological Signs of Ovulation and Fertility Readily Observable by Women. Linacre Q. 2013 Jan 1;80(1):17–23.

11. Su H, Yi Y, Wei T, Chang T, Cheng C. Detection of ovulation, a review of currently available methods. Bioeng Transl Med. 2017 Sep 16;2(3):238–46.

12. Ameyaw EK, Woytowich D, Gbagbo FY, Amoah PA. Assessing geographical variation in ovulatory cycle knowledge among women of reproductive age in Sierra Leone: Analysis of the 2019 Demographic and Health Survey. PLoS One. 2024 Apr 16;19(4):e0300239.

13. Wolde M, Kassie A, Shitu K, Azene ZN. Knowledge of Fertile Period and Its Determinants Among Women of Childbearing age in Ethiopia: A Multilevel Analysis Based on 2016 Ethiopian Demographic and Health Survey. Front Public Health. 2022 May 19;10.

14. Ferrettiid A, Vayena E, Blasimme A. Unlock digital health promotion in LMICs to benefit the youth. 2023; Available from: 10.1371/journal.pdig.0000315

15. Westoff CF, Koffman DA. The Association of Television and Radio with Reproductive Behavior. Popul Dev Rev. 2011 Dec 13;37(4):749–59.

16. Aboagye RG, Seidu AA, Ahinkorah BO, Cadri A, Frimpong JB, Hagan JE, et al. Association between frequency of mass media exposure and maternal health care service utilization among women in sub-Saharan Africa: Implications for tailored health communication and education. PLoS One. 2022 Sep 29;17(9):e0275202.

17. Chipako I, Singhal S, Hollingsworth B. Impact of sexual and reproductive health interventions among young people in sub-Saharan Africa: a scoping review. Front Glob Womens Health. 2024 Apr 18;5.

18. Ghana Statistical Service (GSS), ICF. Ghana Demographic and Health Survey 2022. Accra, Ghana, and Rockville, Maryland, USA; 2024.

19. GSS, ICF. 2022 Ghana Demographic and Health Survey. 2022.

20. Afrifa-Anane GF. Knowledge of the ovulatory cycle and its determinants among adolescent females in Ghana. BMC Womens Health. 2025 Jan 23;25(1):33.

21. Jean Simon D, Jamali Y, Olorunsaiye CZ, Théodat JM. Knowledge of the ovulatory cycle and its determinants among women of childbearing age in Haiti: a population-based study using the 2016/2017 Haitian Demographic Health Survey. BMC Womens Health. 2023 Jan 2;23(1):2.

22. Ross WR, Kiprotich E, Rade BK, Wolde M, Azene ZN, Kassie A, et al. Knowledge of Fertile Period and Its Determinants Among Women of Childbearing age in Ethiopia: A Multilevel Analysis Based on 2016 Ethiopian Demographic and Health Survey. Frontiers in Public Health | www.frontiersin.org [Internet]. 2022;1:828967. Available from: www.frontiersin.org

23. Ayoola AB, Zandee GL, Adams YJ. Women’s Knowledge of Ovulation, the Menstrual Cycle, and Its Associated Reproductive Changes. Birth. 2016 Sep 9;43(3):255–62.

24. Iyanda AE, Dinkins BJ, Osayomi • Tolulope, Temitope •, Adeusi J, Lu Y, et al. Fertility knowledge, contraceptive use and unintentional pregnancy in 29 African countries: a cross-sectional study. 2020; Available from: 10.1007/s00038-020-01356-9

25. Iyanda AE, Dinkins BJ, Osayomi T, Adeusi TJ, Lu Y, Oppong JR. Fertility knowledge, contraceptive use and unintentional pregnancy in 29 African countries: a cross-sectional study. Int J Public Health. 2020 May 9;65(4):445–55.

26. Dagnew B, Teshale AB, Dagne H, Diress M, Tesema GA, Dewau R, et al. Individual and community-level determinants of knowledge of ovulatory cycle among women of childbearing age in Ethiopia: A multilevel analysis based on 2016 Ethiopian Demographic and Health Survey. PLoS One. 2021 Sep 1;16(9 September).

27. Ameyaw EK, Woytowich D, Gbagbo FY, Amoah PA. Assessing geographical variation in ovulatory cycle knowledge among women of reproductive age in Sierra Leone: Analysis of the 2019 Demographic and Health Survey. PLoS One. 2024 Apr 16;19(4):e0300239.

28. Uzoechi CA, Davod Parsa A, Mahmud I, Alasqah I, Kabir R. medicina Menstruation among In-School Adolescent Girls and Its Literacy and Practices in Nigeria: A Systematic Review. 2023; Available from: 10.3390/medicina59122073

29. Msovela J, Shija AE, Ntuyeko H, Imeda C, Mugula A, Mgina E, et al. Puberty and Menstruation Knowledge, Information Sources and Needs among Secondary School Adolescent Girls and Boys in Kibaha, Tanzania. 2025; Available from: 10.1371/journal.pgph.0004176

30. Mandiwa C, Namondwe B, Makwinja A, Zamawe C. Factors associated with contraceptive use among young women in Malawi: analysis of the 2015-16 Malawi demographic and health survey data. 2018; Available from: 10.1186/s40834-018-0065-x

